# Bacterial pulmonary superinfections are associated with unfavourable outcomes in critically ill COVID-19 patients

**DOI:** 10.1101/2020.09.10.20191882

**Authors:** Philipp K. Buehler, Annelies S. Zinkernagel, Daniel A. Hofmaenner, Pedro David Wendel García, Claudio T. Acevedo, Alejandro Gómez-Mejia, Srikanth Mairpady Shambat, Federica Andreoni, Martina A. Maibach, Jan Bartussek, Matthias P. Hilty, Pascal M. Frey, Reto A. Schuepbach, Silvio D. Brugger

**Author notes:** These authors contributed equally to the work (Philipp K. Buehler, Annelies S. Zinkernagel, Daniel A. Hofmaenner). These authors contributed equally to the work (Reto A. Schuepbach, Silvio D. Brugger). Correspondence to: Silvio D. Brugger, M.D., Ph.D., Division of Infectious Diseases and Hospital Epidemiology, University Hospital Zurich, Raemistrasse 100, CH-8091 Zurich, Switzerland.

## Abstract

**Objectives:** While superinfections are associated with unfavourable disease course, their impact on clinical outcomes in critically ill COVID-19 patients remains largely unknown. We aimed to investigate the burden of superinfections in COVID-19 patients.

**Methods:** In this prospective single centre cohort study in an intensive care setting patients aged ≥ 18 years with COVID-19 acute respiratory distress syndrome were assessed for concomitant microbial infections by longitudinal analysis of tracheobronchial secretions, bronchoalveolar lavages and blood. Our primary outcome was ventilator-free survival on day 28 in patients with and without clinically relevant superinfection. Further outcomes included the association of superinfection with ICU length of stay, incidence of bacteremia, viral reactivations, and fungal colonization.

**Results:** In 45 critically ill COVID-19 patients, we identified 19 patients with superinfections (42.2%) by longitudinal analysis of 433 TBS, 35 BAL and 455 blood samples, respectively. On average, superinfections were detected on day 10 after ICU admission. The most frequently isolated clinically relevant bacteria were Enterobacteriaceae, *Streptococcus pneumoniae*, and *Pseudomonas aeruginosa*.

Ventilator-free survival was substantially lower in patients with superinfection (subhazard ratio 0.37, 95%-CI 0.15-0.90, p=0.028). Patients with pulmonary superinfections more often had bacteraemia, virus reactivations, yeast colonization, and needed ICU treatment for a significantly longer time.

**Conclusions:** The detection of superinfections was frequent and associated with reduced ventilator-free survival. Despite empirical antibiotic therapy, superinfections lead to an extended ICU stay in COVID 19 patients. Longitudinal microbiological sampling in COVID-19 patients could allow targeted antimicrobial therapy, and therefore minimize the use of broad-spectrum and reserve antibiotics.

## Introduction

The severe acute respiratory syndrome coronavirus 2 (SARS-CoV-2) evolved as the most relevant pandemic of modern history, challenging health care systems all over the world. The clinical characteristics of coronavirus disease (COVID-19) patients have been thoroughly described in recent studies (1-5). The triggers for acute respiratory distress syndrome (ARDS) in COVID-19 are initially virus-initiated, subsequently leading to inflammation-mediated lung damage and endothelitis (5). Although primarily a viral disease, antibiotics are empirically used in over 70% of cases in addition to experimental antiviral and immunomodulatory treatments (1, 4-7).

Secondary bacterial and/or fungal infections are a well-described phenomenon in viral illnesses such as influenza and are associated with increased morbidity and mortality in viral ARDS as illustrated during previous pandemics (8). Secondary bacterial infections are typically referred to as superinfections whereas co-infection is mainly used to describe simultaneous virus infection. Both, co- and superinfection have been described in COVID-19 patients (6, 9). Data regarding bacterial superinfections in COVID-19 pneumonia are limited and still emerging (10, 11). A recent systematic review has concluded that the rate of bacterial/fungal superinfections is low arguing against the frequent use of broad-spectrum antimicrobials in patients with COVID-19 (10, 12). Still, there is a lack of knowledge about the frequency and significance of bacterial, fungal and viral concomitant infections in critically ill COVID-19 patients.(12) Additionally, in most studies performed so far, no thorough and systematic sampling for concomitant infections was performed. The high mortality in severely ill COVID-19 patients is thought to be at least in part due to secondary infections in addition to viral replication in the lower respiratory tract leading to severe lung injury and ARDS (6, 13, 14).

Superinfection seems to represent a major risk factor for mortality in COVID-19 patients (7, 15-17). However, the risk of superinfection in mechanically ventilated patients with severe COVID-19 remains poorly described.

Currently, the diagnostic and treatment approach for superinfections remains unclear. Classical criteria for the detection of superinfections are often of limited use in COVID-19 patients. Clinical symptoms are an expression of the underlying disease of COVID-19 and cannot be used to reliably distinguish between patients presenting with or without relevant superinfections. For this reason, several authors have argued in favour of an empirical antibiotic treatment with a focus on streptococci and staphylococci in severe courses (14). Other opinion leaders recommend longitudinal sampling of severely ill patients for early detection and treatment during the entire course of the disease (6, 18).

Rapid diagnosis of co- and superinfection may not only help to improve survival but would also allow targeted antimicrobial therapy, improving antimicrobial stewardship throughout the course of the pandemic (18, 19).

The aim of our study was to assess the burden of superinfections and the association with clinical outcomes in critically ill patients with COVID ARDS (CARDS) in a tertiary care ICU with highly regulated antibiotic prescription (20).

## Methods

### Study design and population

This study was performed within the MicrobiotaCOVID cohort study. The MicrobiotaCOVID study is a single-centre, prospective observational study conducted at the Institute of Intensive Care Medicine of the University Hospital Zurich (Zurich, Switzerland) registered at clinicaltrials.gov (ClinicalTrials.gov Identifier: NCT04410263).

Patients with confirmed SARS-CoV-2 infection and CARDS requiring ICU support and mainly invasive mechanical ventilation hospitalized between April 2020 and June 2020 during the first COVID-19 wave in Switzerland were eligible.

Inclusion criteria were age >18 years, SARS-CoV-2 infection as determined by real-time reverse transcriptase-polymerase chain reaction (RT-PCR) positivity of nasopharyngeal and/or pharyngeal swabs, TBS or BAL and hospitalization in the ICU for moderate or severe ARDS according to the Berlin criteria (21).

Exclusion criteria were patients or relatives denying informed consent and patients still being treated in the ICU when the study period ended.

### Ethics and study registration

The study was approved by the local ethics committee of the Canton of Zurich, Switzerland (Kantonale Ethikkommission Zurich BASEC ID 2020 - 00646).

### Study outcomes

The primary outcome was ventilator-free survival on day 28. Secondary outcomes were length of hospital stay, ICU stay and duration of ventilation. Further outcomes included the association of pulmonary superinfection and bacteraemia, other virus co-infections, colonisation with yeast, bacterial infections with multidrug resistance (MDR), and longitudinal laboratory inflammation parameters.

### Sample collection, processing and testing

If the respiratory situation allowed bronchoscopy, BAL was collected upon ICU admission, and during the later course of the disease if medically indicated and upon discretion of the treating physicians. TBS was collected from each ventilated patient at least on day 0 (i.e. upon ICU admission), day 1, day 2, day 3, day 5 and henceforth every 5 days. If the clinical situation did not allow TBS collection, no sampling was performed.

Samples were processed at the Institute for Medical Microbiology and at the Institute for Medical Virology of the University of Zurich. Standard clinical microbiology techniques were used for culturing, isolation and identification of bacterial and fungal microorganisms as previously described (22). SARS-CoV-2 was detected by real-time RT-PCR as previously described (23).

At admission, multiplex PCR for respiratory syncytial virus (RSV) A/B, influenza A/B, adenovirus, coronavirus 229E, HKU1, NL63 and OC43, human bocavirus, human metapneumovirus (hMPV), rhino/enterovirus and parainfluenzavirus 1-4 was performed in nasopharyngeal swabs. Multiplex PCR for the detection of atypical respiratory bacteria was performed on pharyngeal swabs at ICU admission.

Moreover, we assessed serum detection and virus load of the following viruses: herpesviridae (herpes simplex type I and II (HSV 1 and 2), cytomegalovirus (CMV), Epstein-Barr virus (EBV) and human herpes virus 6 (HHV6). Additional virus diagnostics, blood and urine cultures were initiated by the treating physicians according to the clinical situation.

### Data collection and covariates

Clinical and laboratory data were obtained from electronic health records and included demographics, comorbidities / risk factors, medication, ICU scores, laboratory values, organ failure, need for invasive ventilation, need for extracorporeal life support (ECLS), rescue therapies, length of ICU/hospital stay, and experimental therapy (steroids, hydroxychloroquine, lopinavir/ritonavir, remdesivir, tocilizumab and empiric antibiotic therapy).

Daily measurements of inflammatory parameters (CRP/PCT), the leukocyte count and the neutrophil/lymphocyte ratio were routinely performed over the first 16 days after ICU admission.

### Definition of a clinically relevant microorganism in respiratory specimens (TBS and BAL) as a proxy for superinfection

The isolation of clinically relevant microorganisms from respiratory specimen cultures of critically ill COVID-19 patients (TBS and/or BAL) was used as a surrogate parameter for superinfection in concordance with clinical and radiological data. Patients were considered positive for clinically relevant microorganisms if the following pathogens were detected in TBS or BAL: *Streptococcus pneumoniae, Staphylococcus aureus, Klebsiella* spp., *Haemophilus influenzae, Escherichia coli, Enterobacter, Citrobacter* spp., *Pseudomonas aeruginosa*, and *Aspergillus* spp (24).

Consultants of the infectious disease department unrelated to the study group were involved daily in the diagnosis and management of patients including specific antimicrobial treatment. Organisms with low pathogenicity for lung infections such as *Enterococcus* spp., *Candida* spp., coagulase-negative Staphylococci and non-pneumococcal Streptococci were reported but not considered a relevant clinical pathogen of the airways in accordance with the literature (25).

Detection of the herpesviridae herpes simplex 1 and 2 as well as cytomegalovirus, Epstein-Barr virus, human herpes virus 6 in blood were also reported but were considered to be reactivations. Viral co-infections/reactivations were only diagnosed if clinical signs of tracheitis or pathological signs of viral co-infection in cytology were observed.

### Statistical analyses

Due to the unknown rate of concomitant infections in severely ill COVID-19 patients a power calculation was not feasible. Comparisons of population characteristics were performed using Mann-Whitney-U tests and the chi-squared/Fisher exact test for categorical variables, as appropriate. For longitudinal analysis of laboratory parameters, differences between time points and superinfection status were tested using linear mixed effects models. To estimate the effect of superinfections on 28-day ventilator-free survival, we used a competing risk regression model according to Fine & Gray censored at 28 days, with the event of extubation as outcome event and death as the competing risk. An alpha level of 0.05 was considered statistically significant. Statistical analysis was performed using STATA version 15 (StataCorp, College Station, TX), R version 3.6.3 (r-project.org/), SPSS Version 23 (SPSS Science, Chicago, IL, USA) and Graphpad Prism 7 (San Diego, CA, USA).

## Result

### Cohort characteristics

A total of 48 critically ill COVID-19 patients with ARDS were screened in ICU at the University Hospital Zurich between April and June 2020. Three patients had to be excluded from the analysis because patients or relatives denied informed consent (Figure 1). 45 patients with a median age of 60 (54-69) years were included in this study. Most of them were male (35/45, 77.8%). Of the 45 patients, 19 (42.2%) were diagnosed with a superinfection. The median of ventilation duration was 15 days and length of ICU stay was 14 days overall. The median length of hospital stay was 24 days. Baseline characteristics are summarised in Table 1. In general, both groups of patients with and without superinfections were similar with regards to demographics and clinical characteristics. In particular, there were no differences in the severity of the disease and organ dysfunction as assessed by SAPS II and SOFA scores. Intensive care rescue therapies such as prone position (42% vs 90%) and tracheotomy (23% vs 74%) were required more frequently and/or for longer periods in the superinfection group (Table 1).

**Table 1.** Demographic and clinical characteristics as well as risk factors of COVID-19 patients stratified according to presence or absence of relevant pathogens in tracheobronchial secretions (TBS) reflecting superinfection. The data are presented as median (IQR) or number and (percentage %). The two groups were compared using Chi-Square Test/Fisher Exact method for nominal distributed data or the Mann Whitney test for scale level distributed data. The significance level is p <0.05.

|  | Overall (n=45) | No Superinfection (n=26) | Superinfection (n=19) | P-value |
| --- | --- | --- | --- | --- |
| **Baseline Characteristics** |  |  |  |  |
| Age median (IQR) in years | 60 (54-69) | 61.5 (54-71) | 59 (54-69) | 0.654 |
| Male [n/%] | 35 (77.8%) | 19 (73.1%) | 16 (84.2%) | 0.375 |
| Weight [kg] | 83 (75-99) | 80 (72.5-90) | 92 (78-100) | 0.049 |
| Height [cm] | 175 (165-182) | 172 (160-180) | 176 (169-185) | 0.112 |
| Body mass index [kg/m<sup>2</sup>] | 27.8 (25.7-31.6) | 27.5 (25.6-30.6) | 27.8 (26.8-35) | 0.346 |
| **Comorbid conditions** |  |  |  |  |
| Myocardial Infarction / Ischemic Heart Disease | 6 (13.3%) | 5 (19.2%) | 1 (5.3%) | 0.222 |
| Art Hypertension | 26 (57.8%) | 16 (61.5%) | 10 (52.6%) | 0.550 |
| Chronic kidney disease | 12 (26.7%) | 10 (38.5%) | 2 (10.5%) | 0.047 |
| Dialysis | 1 (2.2%) | 1 (3.8%) | 0 (0%) | 0.999 |
| Oral hypoglycaemic agents controlled diabetes | 5 (11.1%) | 3 (11.5%) | 2 (10.5%) | 0.999 |
| Insulin-dependent diabetes | 14 (31.1%) | 7 (26.9%) | 7 (36.8%) | 0.528 |
| Asthma | 1 (2.2%) | 1 (3.8%) | 0 (0%) | 0.999 |
| COPD | 5 (11.1%) | 4 (15.4%) | 1 (5.3%) | 0.378 |
| Cardiovascular disease | 7 (15.6%) | 6 (23.1%) | 1 (5.3%) | 0.211 |
| Renal or liver transplantation | 5 (11.1%) | 4 (15.4%) | 1 (5.3%) | 0.378 |
| Immunosuppression | 7 (15.6%) | 5 (19.2%) | 2 (10.5%) | 0.222 |
| Tumour disease | 4 (8.9%) | 2 (7.7%) | 2 (10.5%) | 0.999 |
| Smoking | 14 (31.1%) | 7 (26.9%) | 7 (36.8%) | 0.625 |
| Alcohol abuse | 1 (2.2%) | 0 (0%) | 1 (5.3%) | 0.999 |
| Drug abuse | 0 (0%) | 0 (0%) | 0 (0%) | 1.000 |

| Medical therapy before admission |  |  |  |  |
| --- | --- | --- | --- | --- |
| Statins | 10 (22.7%) | 6 (24%) | 4 (21.1%) | 0.999 |
| ACE-Inhibitor | 7 (15.9%) | 4 (16%) | 3 (15.8%) | 0.999 |

| Scores/Index |  |  |  |  |
| --- | --- | --- | --- | --- |
| Sepsis-related organ failure assessment score (SOFA) | 8 (5-10) | 8 (3-10) | 9 (7-10) | 0.480 |
| Simplified Acute Physiology Score (SAPS) II | 36 (25-50) | 32.5 (24-50) | 42 (28-51) | 0.275 |
| PaO2/FiO2 ratio low at admission | 122 (94-177) | 129 (97-200) | 108 (85-163.5) | 0.228 |

| Organ failure during ICU stay |  |  |  |  |
| --- | --- | --- | --- | --- |
| Acute kidney disease | 27 (60%) | 15 (57.7%) | 12 (63.2%) | 0.712 |
| Dialysis | 18 (40%) | 9 (34.6%) | 9 (47.4%) | 0.388 |
| Ventilation Therapy | 40 (88.9%) | 21 (80.8%) | 19 (100%) | **0.043** |
| Extracorporeal Life Support (ECLS) | 8 (17.8%) | 3 (11.5%) | 5 (26.3%) | 0.253 |

| Rescue therapies |  |  |  |  |
| --- | --- | --- | --- | --- |
| Prone Position | 28 (62.2%) | 11 (42.3%) | 17 (89.5%) | **0.001** |
| iNO | 11 (24.4%) | 5 (19.2%) | 6 (31.6%) | 0.341 |
| Tracheotomy | 20 (44.4%) | 6 (23.1%) | 14 (73.7%) | **0.001** |

| Timing [days] |  |  |  |  |
| --- | --- | --- | --- | --- |
| Time to ICU admission | 2 (1-6) | 2 (1-6) | 3 (1-6) | 0.636 |
| Duration prone position | 6 (1-10) | 1.5 (1-7) | 6 (4-10) | **0.022** |
| Duration iNO therapy | 3 (1-6) | 1 (1-3) | 4.5 (1-22) | 0.349 |
| Duration Intubation to tracheotomy | 20 (10-33) | 10.5 (9-21) | 27 (15-33) | 0.114 |

| Laboratory values at admission |  |  |  |  |
| --- | --- | --- | --- | --- |
| WBC count (G/L) | 7.7 (5.7-10.7) | 7.5 (5.7-10) | 8 (5.3-13.4) | 0.515 |
| Haemoglobin (gr/l) | 118.5 (101.5-133) | 117 (107-132) | 126 (98-134) | 0.896 |
| Haematocrit (%) | 0.4 (0.3-0.4) | 0.4 (0.3-0.4) | 0.4 (0.3-0.4) | 0.619 |
| Platelet count (G/L) | 199 (169.5-272) | 200 (177-271) | 190 (154-297) | 0.776 |
| ALT(U/L) | 42.5 (25.5-65.5) | 31 (24-60) | 56 (33-72) | **0.008** |
| LDH (U/L) | 676 (527-842.5) | 619 (471-742) | 772 (626-876) | 0.144 |
| Alkaline Phosphatase (U/L) | 74.5 (53.5-103.5) | 77.5 (56-108) | 61 (53-98) | 0.308 |
| Urea (mmol/l) | 6 (4.3-10.3) | 6.2 (4.1-7.9) | 6 (4.4-12.2) | 0.651 |
| Creatinine (umol/l) | 92.5 (67-138.5) | 95 (70-128) | 91 (57-149) | 0.387 |
| Serum Sodium (mmol/L) | 138 (134-141) | 137 (135-140) | 141 (134-146) | 0.203 |
| Serum potassium (mmol/L) | 3.9 (3.7-4.5) | 4.1 (3.7-4.4) | 3.8 (3.5-4.7) | 0.601 |
| CRP (mg/L) | 168.5 (83.5-276.5) | 124 (62-238) | 255 (102-301) | **0.034** |
| PCT (mcg/L) | 0.3 (0.2-1.2) | 0.2 (0.1-1.7) | 0.4 (0.2-1.2) | 0.060 |
| IL6 (ng/L) | 127 (71.2-454) | 122 (84-697) | 127 (62.7-263) | 0.554 |

| Special drug therapy |  |  |  |  |
| --- | --- | --- | --- | --- |
| Steroids | 21 (46.7%) | 8 (30.8%) | 13 (68.4%) | **0.012** |
| Hydroxychloroquine | 27 (61.4%) | 13 (52%) | 14 (73.7%) | 0.143 |
| Lopinavir/ritonavir | 7 (15.9%) | 4 (16%) | 3 (15.8%) | 0.999 |
| Remdesivir | 8 (18.2%) | 4 (16%) | 4 (21.1%) | 0.704 |
| Tocilizumab | 2 (4.7%) | 2 (8.3%) | 0 (0%) | 0.501 |
| empiric antimicrobial therapy | 40 (88.39%) | 22 (88%) | 18 (94.7%) | 0.441 |

| Sample size overall |  |  |  |
| --- | --- | --- | --- |
| TBS | 433 | 114 | 319 |
| BAL | 35 | 12 | 23 |
| Blood culture | 455 | 152 | 303 |

**Figure 1.**
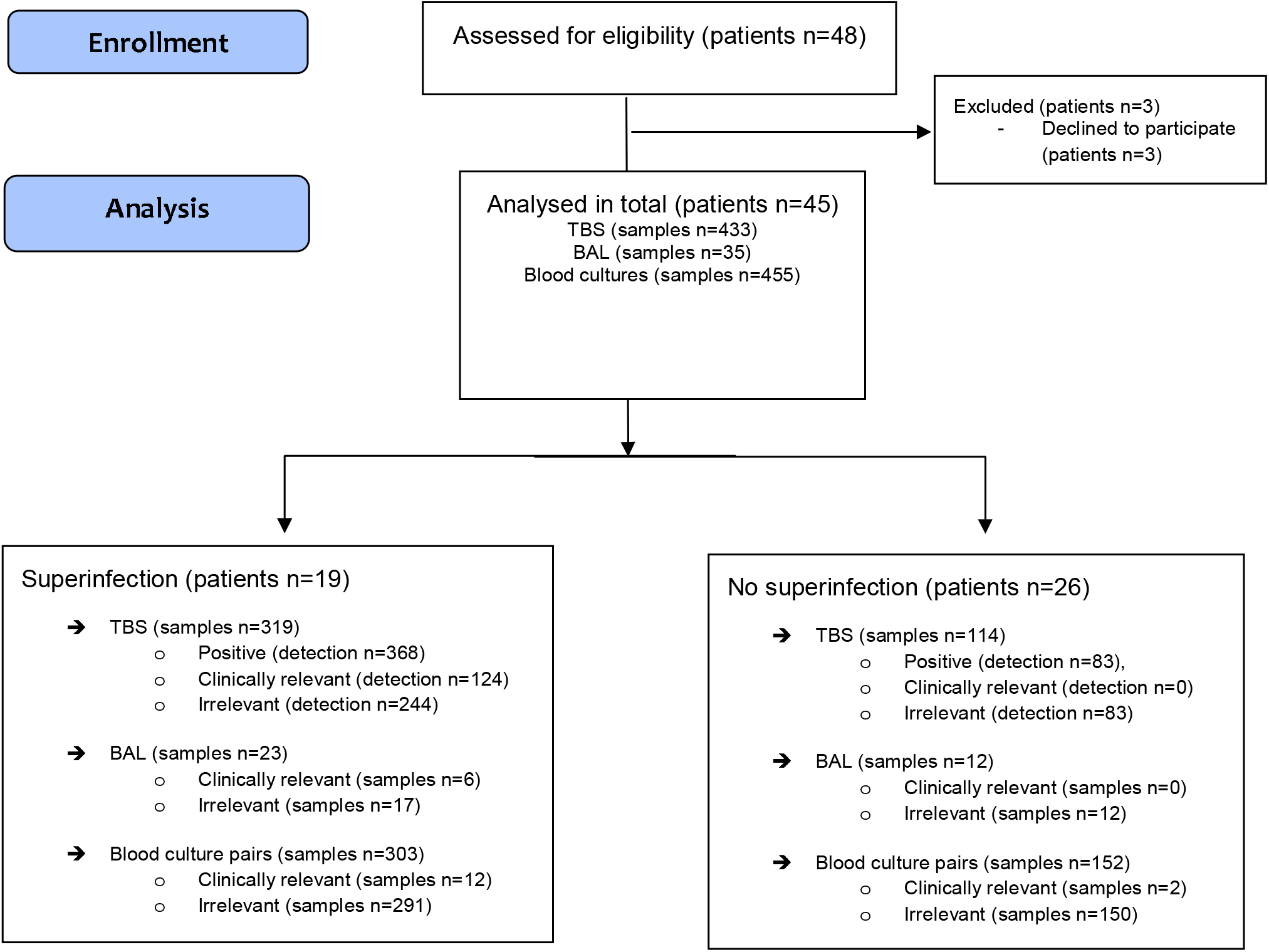
Study Flow Diagram.

### Microbiological sampling and superinfections

Overall, 433 TBS, 35 BAL samples and 455 blood culture pairs were analysed for the presence of relevant microorganisms (Figure 1).

Clinically relevant microorganisms were detected in TBS in almost half the cases. In nineteen patients (42.2%) at least one clinically relevant bacterium or fungus was detected in TBS during the study period, whereas in 26 patients (57.8%) no relevant microorganisms were detected in TBS.

A total of 319 TBS were collected in the superinfection group and 124 relevant microorganisms were detected in these samples. Furthermore, relevant pathogens were detected six times in 23 BAL samples (26.1%). Only in two cases relevant results did not match between BAL and TBS. Despite high frequency of positive TBS, blood cultures showed only seven different bacterial species in twelve positive blood culture pairs (Figure 1).

In the group without superinfections, 83/114 TBS samples showed growth but without recovery of clinically relevant lung pathogens. *Candida albicans* was the most frequently isolated non-relevant organism. In BAL there was also no evidence of relevant microorganisms in the 12 9samples. However, clinically relevant bacteraemia was detected twice in a total of 152 blood culture pairs (Figure 1).

The detection time points of clinically relevant and non-relevant microorganisms are depicted in Figure 2. On average, clinically relevant pathogens were detected on day ten after ICU admission and reflect the hospital-acquired pneumonia (HAP) / ventilator-associated pneumonia (VAP) spectrum (Figure 2A). Non-relevant pathogens were detected on average on day three post ICU admission (Figure 2B). The most frequently isolated bacteria per patient were detected: *Enterococcus* spp. (15/45), *Enterobacter/Citrobacter* (8/45) and *Klebsiella* spp. (7/45). Additionally, *Streptococcus pneumoniae* (2/45), *Escherichia coli* (2/45), *Enterobacter* spp. (5/45), *Citrobacter* spp. (3/45), *Pseudomonas aeruginosa* (5/45), *Burkholderia cepacia* (2/45) and Staphylococci (all coagulase-negative) (13/45)

Empirical antimicrobial therapy was given to 40/45 (88.9%) patients, antifungal therapy to 10/45 (22.2%) patients and antiviral therapy to treat concomitant viral infections to 9/45 (20 %) patients. Figures 2C and D summarize the antibiotic treatment.

In ten patients (22.2%) multi drug resistant (MDR) bacteria were detected *(Pseudomonas aeruginosa, Entereobacter cloacae* and *Burkholderia cepacia)*.

Serum reactivation of HSV 1 and 2 was detected in 5 out of 45 patients. HHV 6 was detected twice, CMV reactivation occurred once and EBV reactivation twice. One patient had a co-infection with influenza A (Table 2).

**Table 2.**
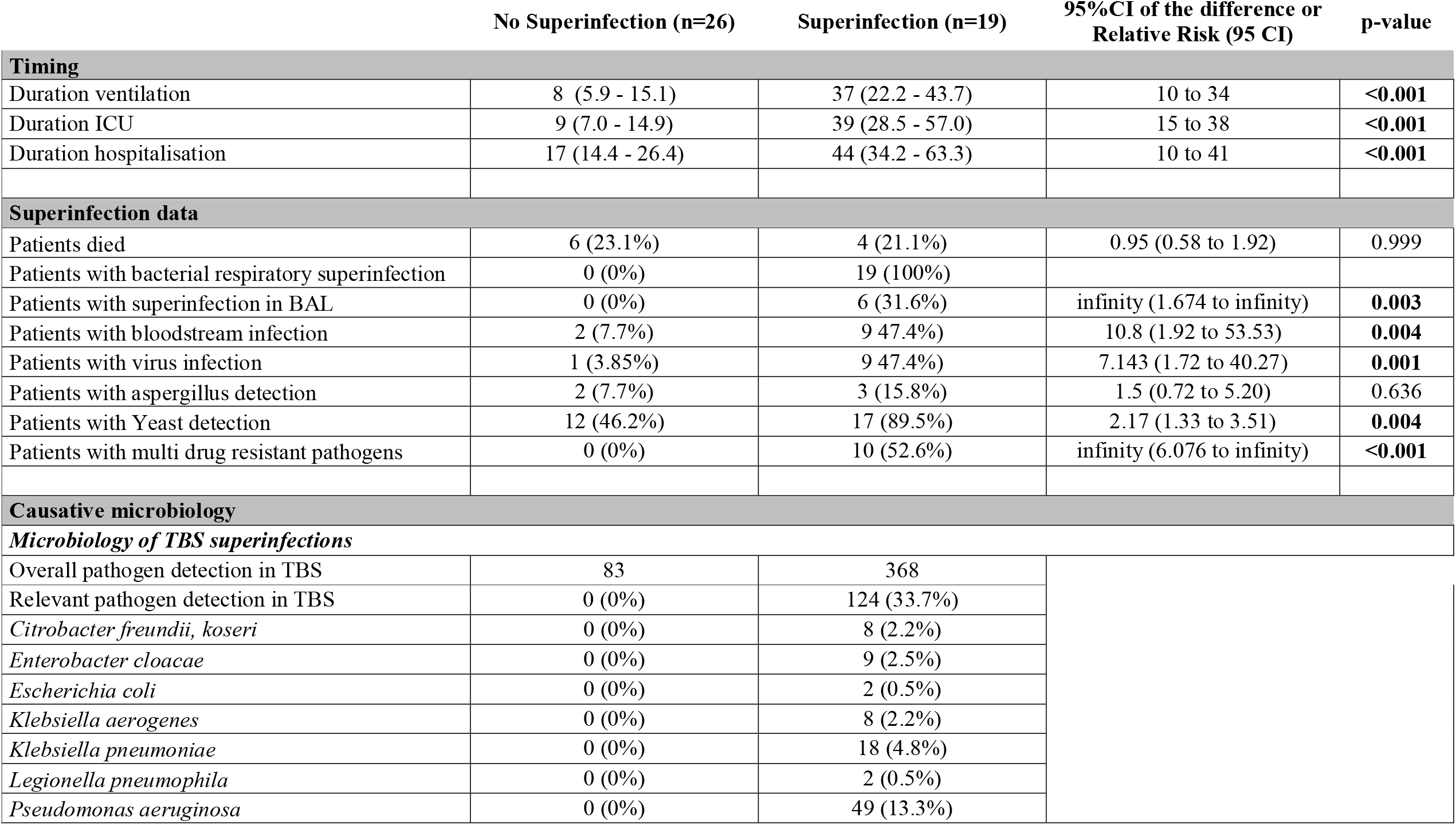
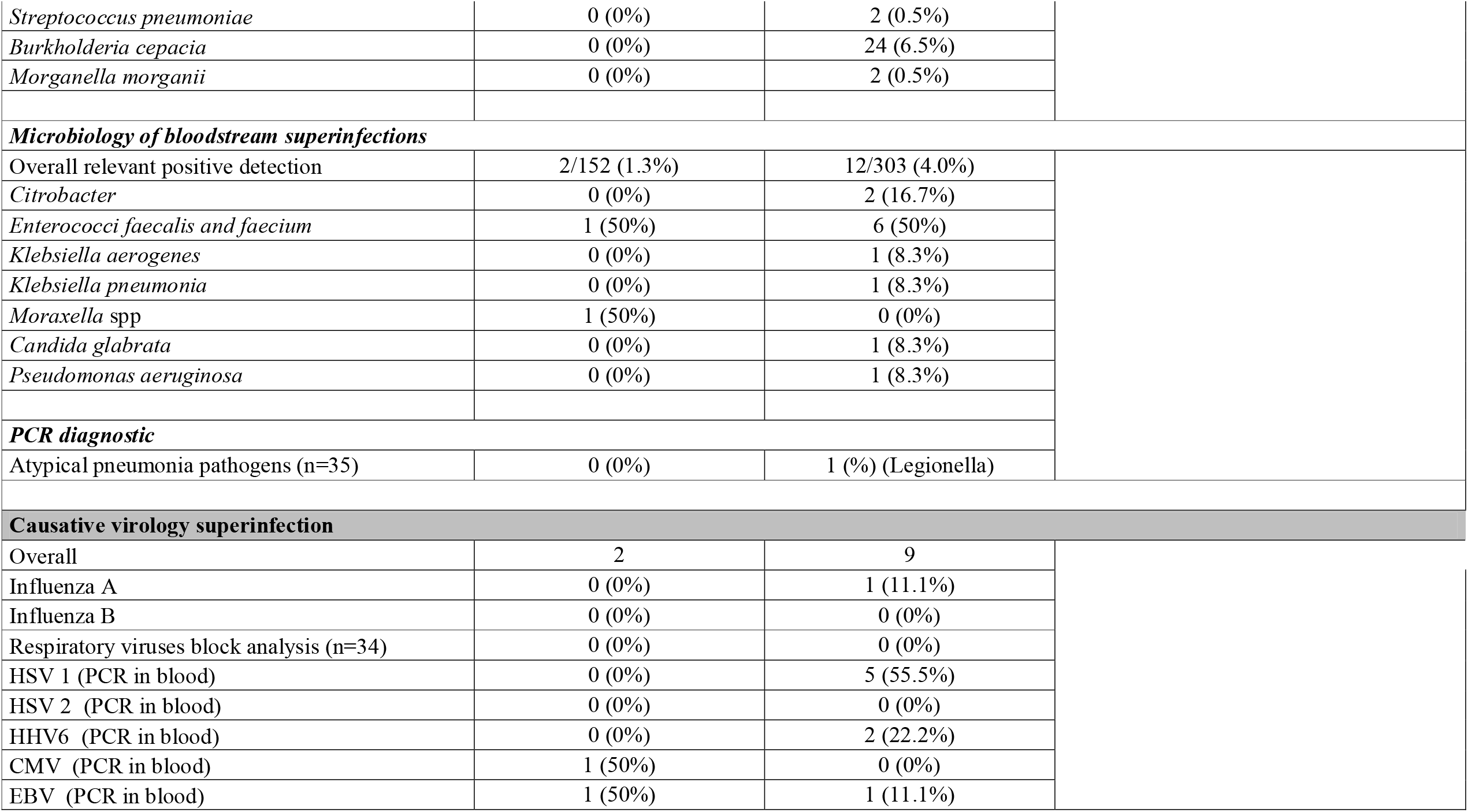
Clinical outcomes and microorganisms detected. The data are presented as median (95 CI of Median) or number and (percentage %). 95%CI of the difference or Relative Risk (95 CI) were calculated too. The two groups were compared using Chi-Square Test/Fisher Exact method for nominal distributed data or the Mann Whitney test for scale level distributed data. The significance level is p <0.05.

|  | No Superinfection (n=26) | Superinfection (n=19) | 95%CI of the difference or Relative Risk (95 CI) | p-value |
| --- | --- | --- | --- | --- |
| **Timing** |  |  |  |  |
| Duration ventilation | 8 (5.9 - 15.1) | 37 (22.2 - 43.7) | 10 to 34 | **<0.001** |
| Duration ICU | 9 (7.0 - 14.9) | 39 (28.5 - 57.0) | 15 to 38 | **<0.001** |
| Duration hospitalisation | 17 (14.4 - 26.4) | 44 (34.2 - 63.3) | 10 to 41 | **<0.001** |
| **Superinfection data** |  |  |  |  |
| Patients died | 6 (23.1%) | 4 (21.1%) | 0.95 (0.58 to 1.92) | 0.999 |
| Patients with bacterial respiratory superinfection | 0 (0%) | 19 (100%) |  |  |
| Patients with superinfection in BAL | 0 (0%) | 6 (31.6%) | infinity (1.674 to infinity) | **0.003** |
| Patients with bloodstream infection | 2 (7.7%) | 9 47.4%) | 10.8 (1.92 to 53.53) | **0.004** |
| Patients with virus infection | 1 (3.85%) | 9 47.4%) | 7.143 (1.72 to 40.27) | **0.001** |
| Patients with aspergillus detection | 2 (7.7%) | 3 (15.8%) | 1.5 (0.72 to 5.20) | 0.636 |
| Patients with Yeast detection | 12 (46.2%) | 17 (89.5%) | 2.17 (1.33 to 3.51) | **0.004** |
| Patients with multi drug resistant pathogens | 0 (0%) | 10 (52.6%) | infinity (6.076 to infinity) | **<0.001** |
| **Causative microbiology** |  |  |  |  |
| **Microbiology of TBS superinfections** |  |  |  |  |
| Overall pathogen detection in TBS | 83 | 368 |  |  |
| Relevant pathogen detection in TBS | 0 (0%) | 124 (33.7%) |  |  |
| *Citrobacter freundii, koseri* | 0 (0%) | 8 (2.2%) |  |  |
| *Enterobacter cloacae* | 0 (0%) | 9 (2.5%) |  |  |
| *Escherichia coli* | 0 (0%) | 2 (0.5%) |  |  |
| *Klebsiella aerogenes* | 0 (0%) | 8 (2.2%) |  |  |
| *Klebsiella pneumoniae* | 0 (0%) | 18 (4.8%) |  |  |
| *Legionella pneumophila* | 0 (0%) | 2 (0.5%) |  |  |
| *Pseudomonas aeruginosa* | 0 (0%) | 49 (13.3%) |  |  |

|  |  |  |
| --- | --- | --- |
| *Streptococcus pneumoniae* | 0 (0%) | 2 (0.5%) |
| *Burkholderia cepacia* | 0 (0%) | 24 (6.5%) |
| *Morganella morganii* | 0 (0%) | 2 (0.5%) |
| **Microbiology of bloodstream superinfections** |  |  |
| Overall relevant positive detection | 2/152 (1.3%) | 12/303 (4.0%) |
| *Citrobacter* | 0 (0%) | 2 (16.7%) |
| *Enterococci faecalis and faecium* | 1 (50%) | 6 (50%) |
| *Klebsiella aerogenes* | 0 (0%) | 1 (8.3%) |
| *Klebsiella pneumonia* | 0 (0%) | 1 (8.3%) |
| *Moraxella* spp | 1 (50%) | 0 (0%) |
| *Candida glabrata* | 0 (0%) | 1 (8.3%) |
| *Pseudomonas aeruginosa* | 0 (0%) | 1 (8.3%) |
| **PCR diagnostic** |  |  |
| Atypical pneumonia pathogens (n=35) | 0 (0%) | 1 (%) (*Legionella*) |
| **Causative virology superinfection** |  |  |
| Overall | 2 | 9 |
| Influenza A | 0 (0%) | 1 (11.1%) |
| Influenza B | 0 (0%) | 0 (0%) |
| Respiratory viruses block analysis (n=34) | 0 (0%) | 0 (0%) |
| HSV 1 (PCR in blood) | 0 (0%) | 5 (55.5%) |
| HSV 2 (PCR in blood) | 0 (0%) | 0 (0%) |
| HHV6 (PCR in blood) | 0 (0%) | 2 (22.2%) |
| CMV (PCR in blood) | 1 (50%) | 0 (0%) |
| EBV (PCR in blood) | 1 (50%) | 1 (11.1%) |

Furthermore, colonisations with fungi were detected and the isolated organisms included *Candida* spp. (29/45 patients), non-*Candida* yeast (21/45 patients) and *Aspergillus* spp. (5/45 patients). A detailed overview of relevant pathogens detected in TBS and blood cultures are shown in Table 2.

### Ventilator-free survival at 28 days

COVID-19 patients with pulmonary superinfections had a substantially lower 28-day ventilator-free survival than those without superinfections (Figure 3), with a subhazard ratio of 0.37 (95% Confidence Interval 0.15 - 0.90, p = 0.028).

### Secondary outcomes

Patients with superinfections detected in respiratory specimens were ventilated for significantly longer time periods (8 vs 37 days, p<0.001) and had a significantly longer duration of stay in the ICU (9 vs 39 days, p<0.001) and overall hospitalization time (17 vs 44 days, p<0.001) as compared to patients without superinfections (Table 2).

### Further outcomes

Patients with pulmonary superinfections had significantly more bacteraemia (p=0.004), virus co-infections other than SARS-CoV-2 (p=0.001), colonisation with yeast (p=0.004) and infections with MDR pathogens (p<0.001) (Table 2).

Longitudinal laboratory inflammation parameters (leukocytes, CRP, PCT and Neutrophil/Lymphocyte ratio) for days 1-16 are shown in Supplemental Figure 1A and Supplemental Table 1. Only for CRP, the mixed model evaluation showed a significant difference with increased CRP in the superinfection group (p <0.001). (Supplemental Figure 1 B, C and D).

## Discussion

In this prospective cohort study of critically ill, ventilated COVID-19 patients the presence of respiratory bacteria as a proxy of superinfection was associated with extended ventilation times, increased duration of intensive care and hospitalization and increased need for intensive-care rescue therapies. Bacterial superinfections were detected in 42.2% of patients in our cohort, which is slightly higher than reported in previous studies (1, 2, 5, 6, 13, 14, 26). This discrepancy with other studies might be mainly due to the nature of our cohort consisting of severely ill patients with CARDS. In addition, differing from other studies, sampling was prospectively and repetitively scheduled and not only performed at admission as in other studies, which may account for under-reporting of superinfections. Regional differences can also play an important role especially in bacterial superinfections and spectrum of resistance. This can explain the increased rate of superinfections compared to the existing meta-analyses (10, 27, 28).

Although some studies have concluded that bacterial superinfections do not play a major role in disease severity and treatment choices, the results of the present study challenges the generalizability to severely ill CARDS patients (6, 10). In our cohort, isolation of relevant respiratory bacteria was associated with more severe COVID-19 disease courses with significantly longer duration of invasive mechanical ventilation and prolonged ICU and hospital stays. Compared to other studies investigating the role of superinfection in COVID-19, duration of ICU stay and length of ventilation was high, reflecting the disease severity of patients included in this study (7, 10). Additionally, data on the duration of ventilation and ICU stay are often missing in other studies making comparisons difficult (4, 10, 13, 29, 30). Furthermore, due to the comparatively high SOFA-Score upon admission but moderate mortality, as in our cohort, long-term ICU-treatment complications such as nosocomial infections become more frequent.

Relevant respiratory bacteria were isolated on average on day ten after ICU admission in our cohort suggesting mainly nosocomial infections. In contrast to bacterial superinfections observed in influenza pneumonia, COVID-19 superinfections with Gram-positive bacteria, such as Pneumococci or Staphylococci, were rare in this study (31). Similar observations were also made for MERS-CoV and SARS-CoV-1 associated superinfections (32-34). In this study, mainly Gram-negative pathogens such as *Pseudomonas* and Enterobacteriaceae including MDR bacteria were isolated, which is in line with previous reported studies (10).

Based on the finding that pulmonary bacterial superinfections seem to be mostly nosocomial, empirical broad-spectrum antimicrobial therapy could be stopped and only treated, if pathogenic bacteria are detected (12, 30). Future prospective, randomized trials to investigate the efficacy of targeted antimicrobial therapy should be conducted to define best practice regarding prevention and treatment of bacterial superinfections in COVD-19. The isolation of mainly Gram-negative rods including MDR led to the use of reserve antibiotics after initial empirical therapy of nosocomial pneumonia (Figure 2). It is important to consider the short and long-term consequences that the use of antimicrobials, especially regarding broad spectrum and reserve antibiotics, may have on drug-resistance. A worrisome potential consequence of the COVID-19 pandemic might be the long-term spread of antimicrobial resistance (AMR) due to increased exposure of patients to antimicrobial agents that may have been used inappropriately (35). In this framework, employment of standardized longitudinal screening with early detection and susceptibility testing before establishment of targeted antimicrobial therapy, could minimize the use of broad-spectrum and reserve antibiotics, thus reducing AMR.

The high rate of yeast detection might be associated with the widespread use of broad-spectrum empirical antimicrobial therapy (36). Invasive aspergillosis was not detected (37, 38). So far, only few studies have investigated fungal superinfections in COVID-19 patients (13, 17, 39). The significance of viral reactivation remains unclear (10). In our study, reactivations of HSV 1, HSV 2 and HHV 6 in the serum occurred in patients with bacterial superinfections. These findings support the hypothesis that superinfections associated with increased COVID-19 disease severity might enhance susceptibility to viral reactivations. Further studies with higher participant numbers should clarify the significance of this finding.

In line with previous studies, conventional clinical laboratory tests such as leukocytes, PCT and neutrophil/lymphocyte ratio progressions were not associated with pulmonary superinfections and therefore do not seem very useful for the detection of bacterial superinfections in COVID-19 patients on mechanical ventilation. This complicates the diagnosis of bacterial superinfections and emphasizes the importance of longitudinal microbiological diagnostics.

Advantages of this study are the prospective longitudinal monitoring of respiratory materials with concomitant recording of demographic data, microbiological evaluations and antimicrobial therapy in a tertiary care centre in a high-resource setting that did not experience health care shortage during the first pandemic wave. Furthermore, this study used strict definitions for relevant respiratory pathogens and the diagnosis of superinfections was performed prospectively on defined days longitudinally with detection not only of bacterial but also fungal and viral super-and co-infections.

Limitations of the study are the single centre design, small number of patients and the high number of patients with empirical broad-spectrum antibiotic therapy (>90% of cases) at admission. Another limitation is the lack of a uniform internationally valid definition of a bacterial infection of the lower respiratory tract.

In summary, the detection of relevant bacterial pulmonary superinfection was associated with a more severe disease course in COVID-19 patients, especially a lower likelihood of ventilator-free survival at 28 days. Future trials should investigate the effect of tailored antimicrobial therapy on outcome, antibiotic resistance and drug use based on longitudinal assessment of respiratory tract cultures.

## Data Availability

All data is available upon request.

## Transparency Declaration

### Authors’ contributions

PKB, ASZ, RAS, SDB designed the study and provided funding and infrastructure. PKB and SDB were responsible for the ethical approval. PKB, PWG, DAH, PMF and SDB performed the statistical analysis. PKB, ASZ, DAH, PDW, CTA, AGM, SMS, FA, MAM, JB, MPH, PMF, RAS and SDB collected, analysed and interpreted the data. PKB, DAH, SDB, ASZ and FA wrote the first draft of the manuscript.

All authors revised the manuscript and approved the final version.

---

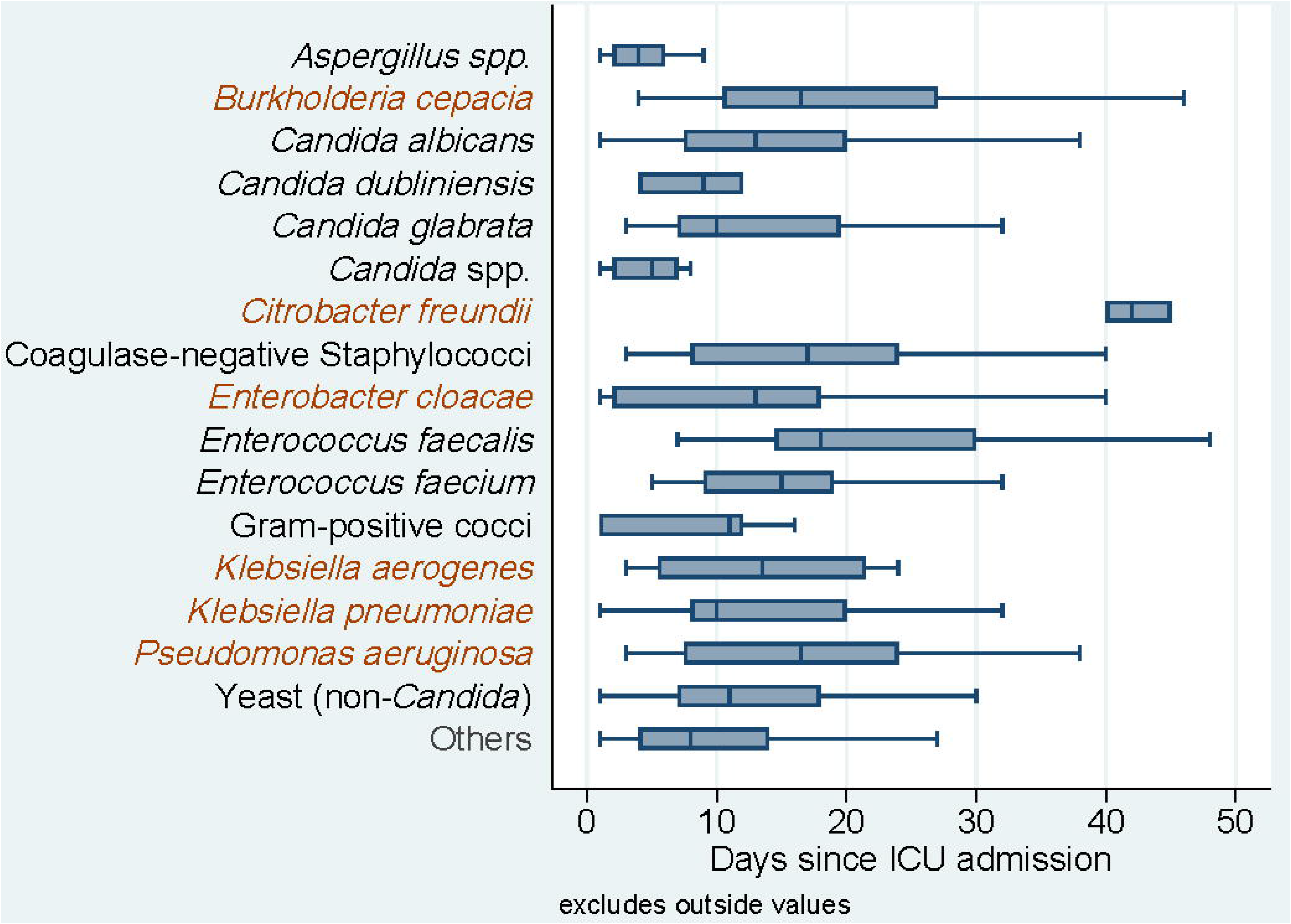

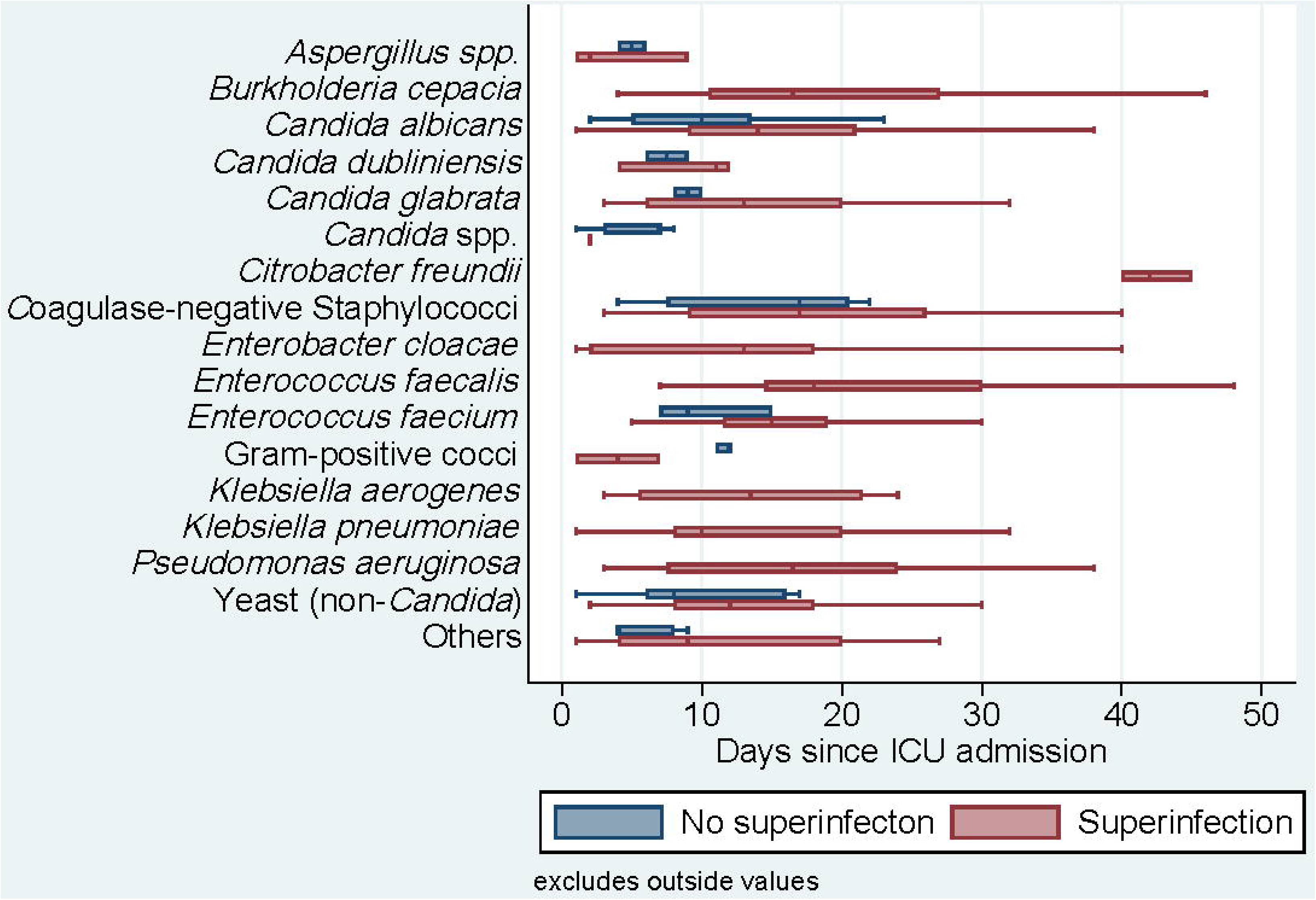

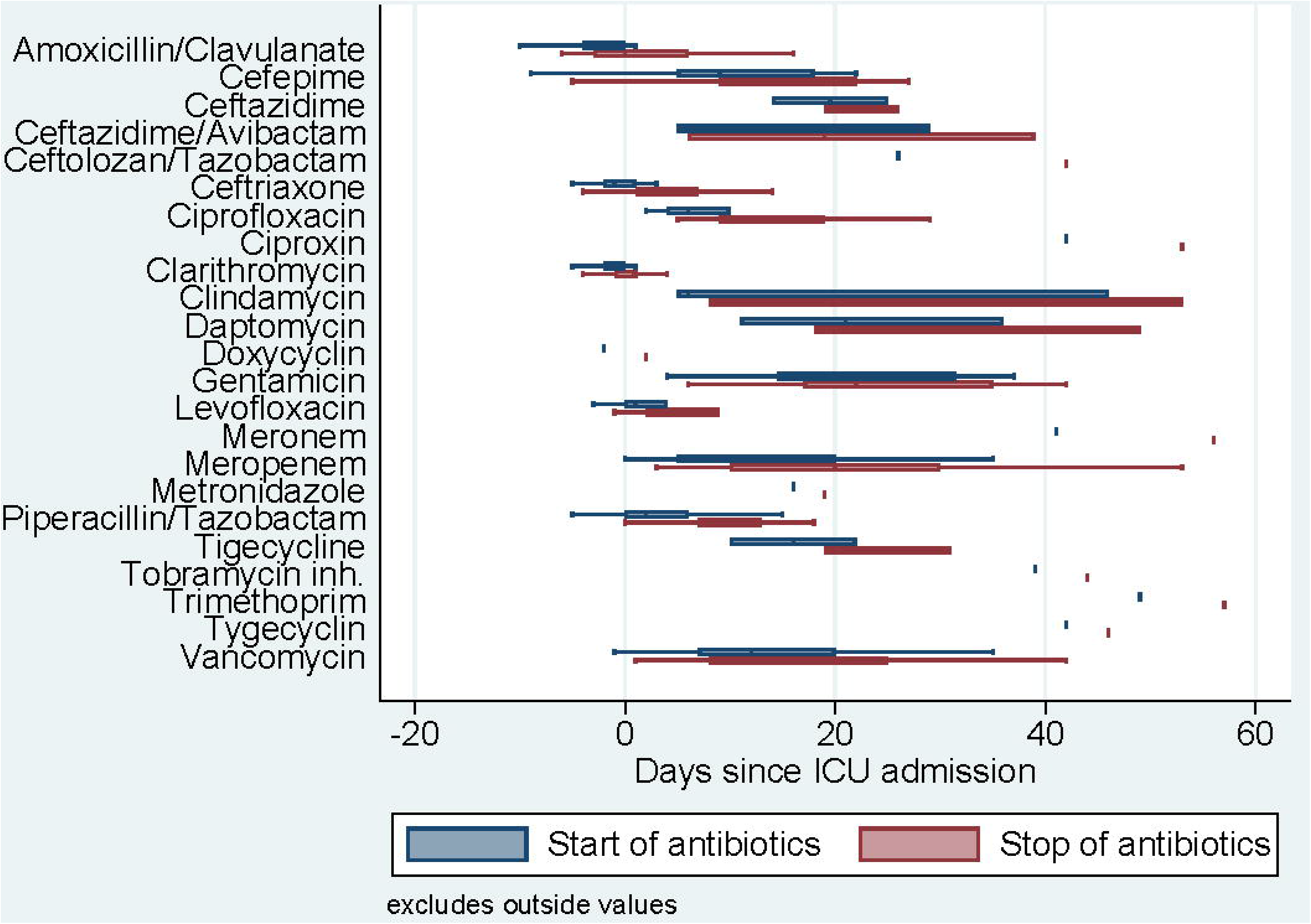

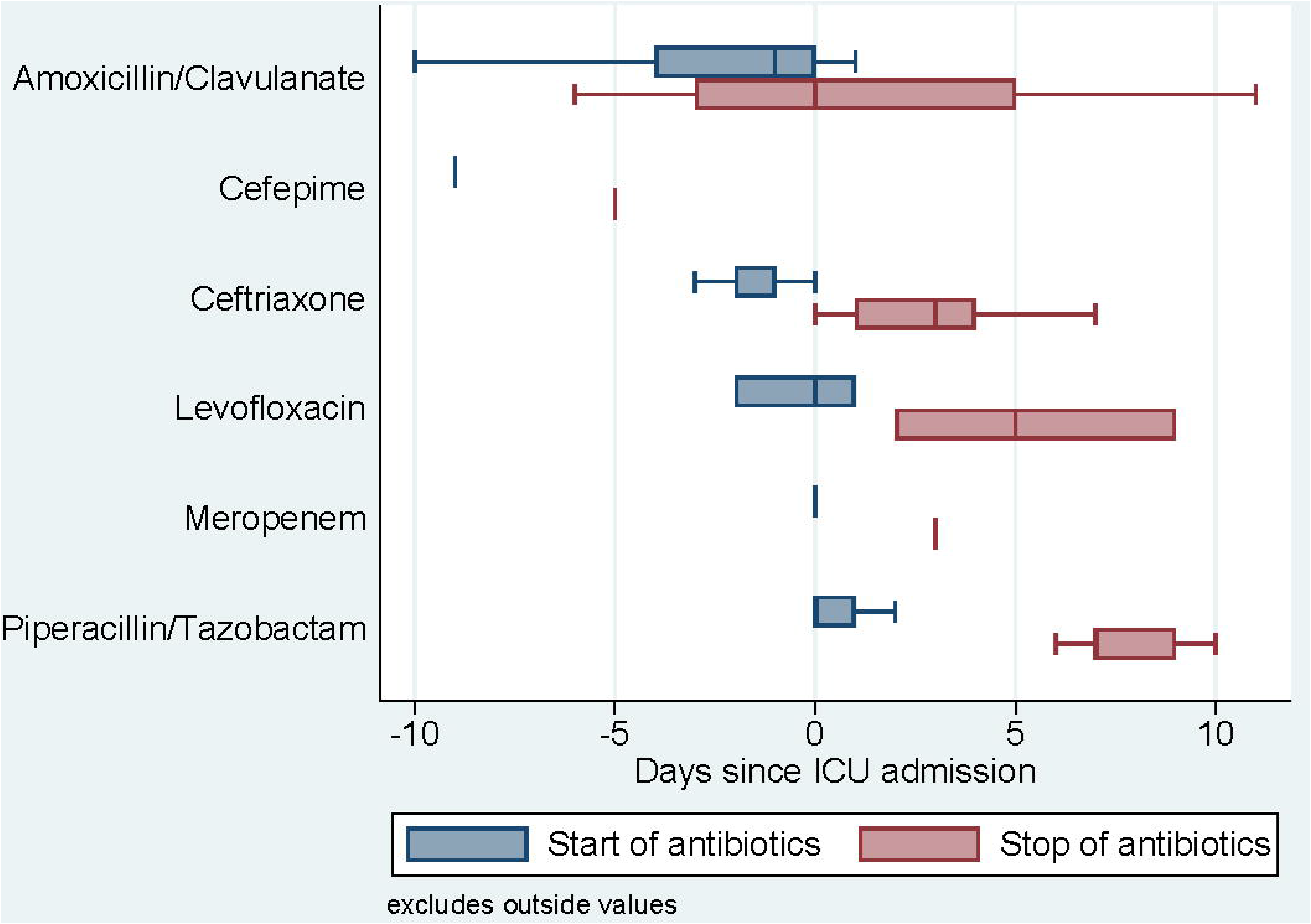

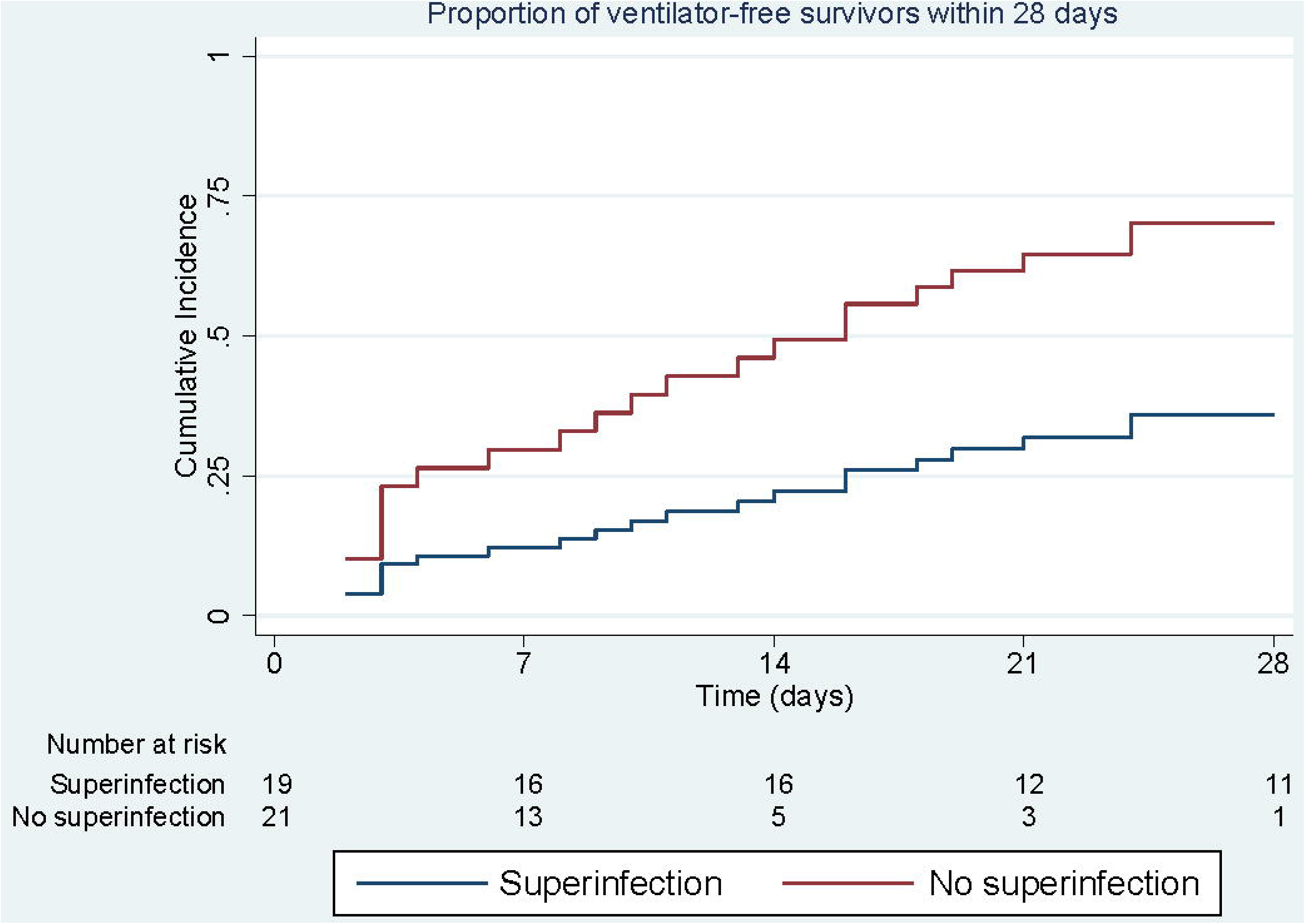

